# Forecasting new daily confirmed cases infected by COVID-19 in Italy from April 9^th^ to May 18^th^ 2020

**DOI:** 10.1101/2020.10.30.20223222

**Authors:** Babak Jamshidi, Amir Talaei-Khoei, Shahriar Jamshidi Zargaran, Mansour Rezaei

## Abstract

We aim at forecasting the outbreak of COVID-19 in Italy by using a two-part time series to model the daily relative increments. Our model is based on the data observed from 22 February to 8 April 2020 and its objective is forecasting 40 days from 9 April to 18 May 2020. All the calculations, simulations, and results in the present paper have been done in MatLab R2015b. The average curve and 80% upper and lower bounds are calculated based on 100 simulations of the fitted models. According to our model, it is expected that by May 18^th^, 2020, the relative increment (RI) falls to the interval of 0.31% to 1.24% (average equal to 0.78%). During the last three days of the studied period, the RI belonged to the interval 2.5% to 3%. Accordingly, It is expected that the new daily confirmed cases face a decreasing to around 1900 on average. Finally, our prediction establishes that the cumulative number of confirmed cases reaches 237635 (with 80% confidence interval equal to [226340 248417] by May 18^th^, 2020.

## Introduction

The COVID-19 is an ongoing pandemic caused by the severe acute respiratory syndrome coronavirus 2 (SARS-CoV-2). It was first identified in Wuhan, Hubei, China in late December 2019. Until 8 April, around 1500,000 cases have been confirmed in around 210 countries, with epicenters China, South Korea, Italy, Iran, France, USA, Germany, Spain, Switzerland, UK, and Turkey, and over 87,000 people have died from the disease [1-2].

During the COVID-19 pandemic, the first confirmed case in Italy happened on 31 January 2020. On 22 February, the first death was reported, and by the beginning of March, the virus had spread to all regions of Italy [1, 3]. Italy accompanied by Iran simultaneously on 1 March 2020, jointed the first two epicenters of COVID-19; China and South Korea. As of April 8^th^, Italy by 17669 deaths, has experienced the most victims worldwide. Italy had gotten the most confirmed cases in Europe until 4 April [2]. On 8 April 2020, Italy has gotten around 140 K confirmed cases.

Over the past few decades, numerous forecasting methods have been presented in the area of epidemic forecasting. Such methods can be categorized into different classes such as deterministic versus probabilistic, comparative versus generative, macro-dynamic versus micro-dynamic, and so on [4]. One of the probabilistic generative methods to study the epidemic data modeling by time series. The time series models have long been of interest in the literature. The applied time series models in this field are restricted to the ARIMA family, for example [5-8]. In 2020, Jamshidi et al [9] introduced a time series not belonging to ARIMA family to model the decreasing relative increment of spreading of an outbreak. Jamshidi et al [10] applied this model to represent the propagation of the pandemic COVID-19 in Iran. On 15 March 2020, they forecasted that the number of the confirmed cases in Iran gets 71000 on average up to 15 April. Considering 66220 confirmed cases until 9 April, It seems that the model works well.

Accordingly, we want to consider the spread of the pandemic in Italy by modelling the relative increment of the confirmed cases by a two-part time series. The first part for representing the propagation of the disease in the first period when the rate of the transition is high, and the second part for the period of decreasing rate which is indicated by an irreversible fall in the relative increment and continues to the extinction of the disease [9]. It is noticeable that we make a change to correct the model to be able to represent the behavior of the disease in the first period much better. Since the relative increment in this period is much more chaotic, we take the variance of the model as double the analogous in the model introduced by [9].

### Model, Estimation, Simulation, Prediction

Let *Y*_*t*_ is a time series of the number of confirmed cases up to time *t*. We aim at studying 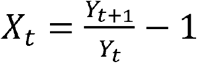, say the daily relative increment. The model we applied has five positive parameters (*b, IR, K, θ, a*);

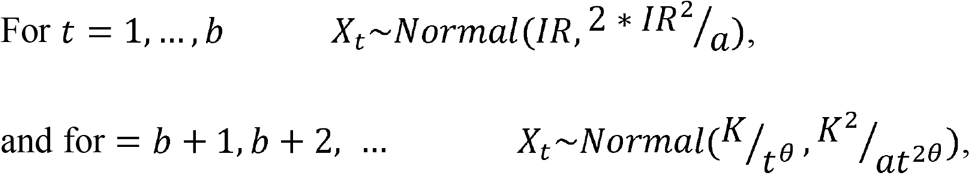

where;

*b*: The length of the first periods of the spread of the disease,

*IR*: The geometric mean of the relative increment in the first periods of the spread of the disease,

*θ*: The acceleration of falling of the relative increments after the first days of spreading (*X*_*t*_ ∝ *t* ^−*θ*^ *for t* = *b* + 1,*b* + 2,…),

*a*: The fixed ratio of the mean to the variance 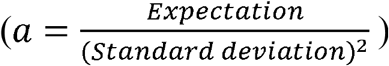, and finally,

*K*: The adjusting coefficient for the curve *t*^−θ^ to fit the time series of the relative increment after the first period of the spread [9].

To estimate the parameters of the model, we can

- the *b* as the first point that the geometric mean of the relative increments in the previous points exceeds 3/2 times the geometric mean of the next three points. Graphically, this time can be identified as the time when the plot of relative increments falls irreversibly (Plot 1). In this case, it is resulted that 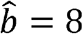.
- calculate the geometric mean of the ratio cumulative numbers in the previous points (1 + *X*_*t*_) from *t* = 1 to *t* = *b* as the estimation of the parameter *IR*.
- Estimate the parameters *θ* and *K* due to the following linear relation

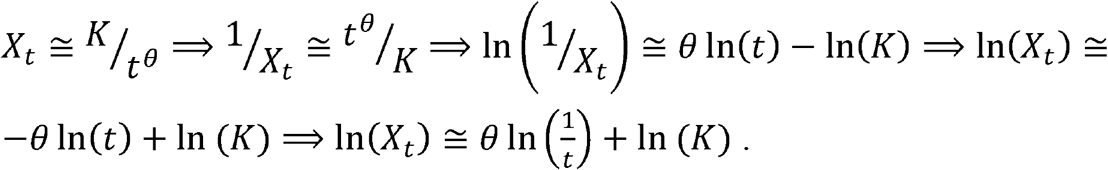

About this dataset, the calculations lead in 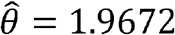 and 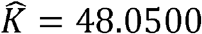 (Plot 2).
- Multiply all the observations after *t* = *b* by 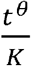 to have an identical mean and variance for all of the newly obtained data (*w*_*t*_)

**Plot 1.**
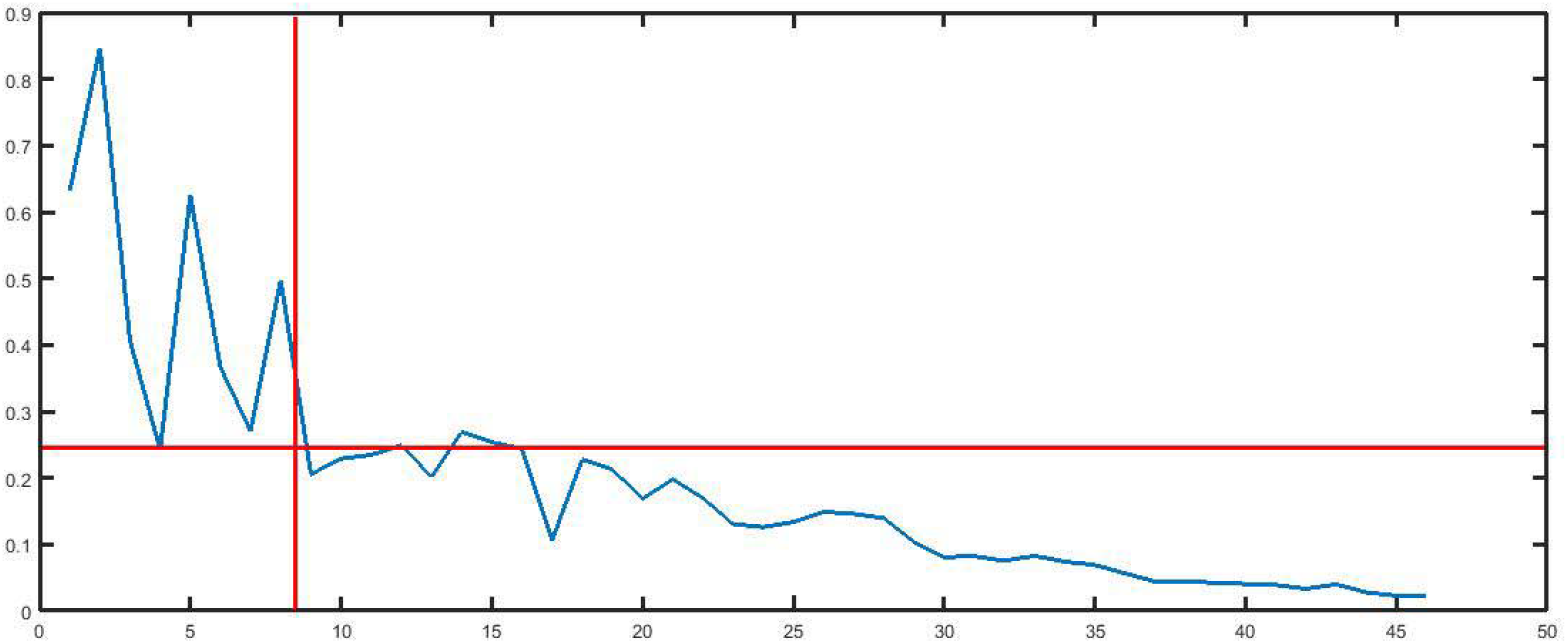
The time series of relative increments and the time of passing from the first stationary period to the new decreasing period.

**Plot 2.**
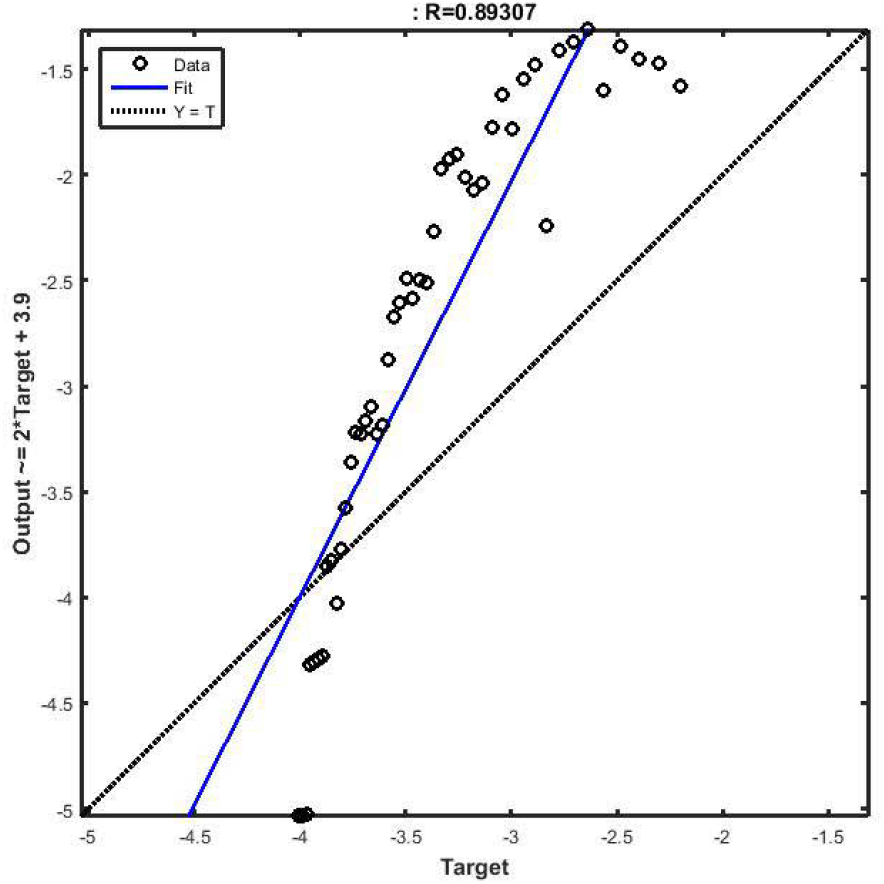
Estimating the linear regression to fit the association between relative increment and the inverse of time to obtain estimations of the parameters *θ* and *K*.

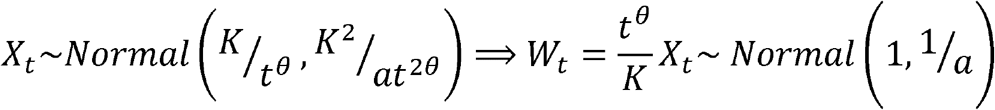

Therefore, the variance of the newly obtained data is a good candidate for estimating 1/*a*. Accordingly, 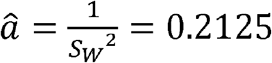.

Overall, we get the following estimations to simulate the model;

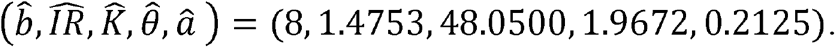

It is worth saying that All the calculations, simulations, and results in our paper have been conducted in MatLab R2015b, and the average curve and 80% upper and lower bounds are calculated based on 100 simulations of the fitted models.

You can see the power of fitting this model to our observations; the relative increments of the cumulative number of confirmed cases infected by COVID-19 in Italy from 23 February to 08 April 2020 (Plot 3).

**Plot 3.**
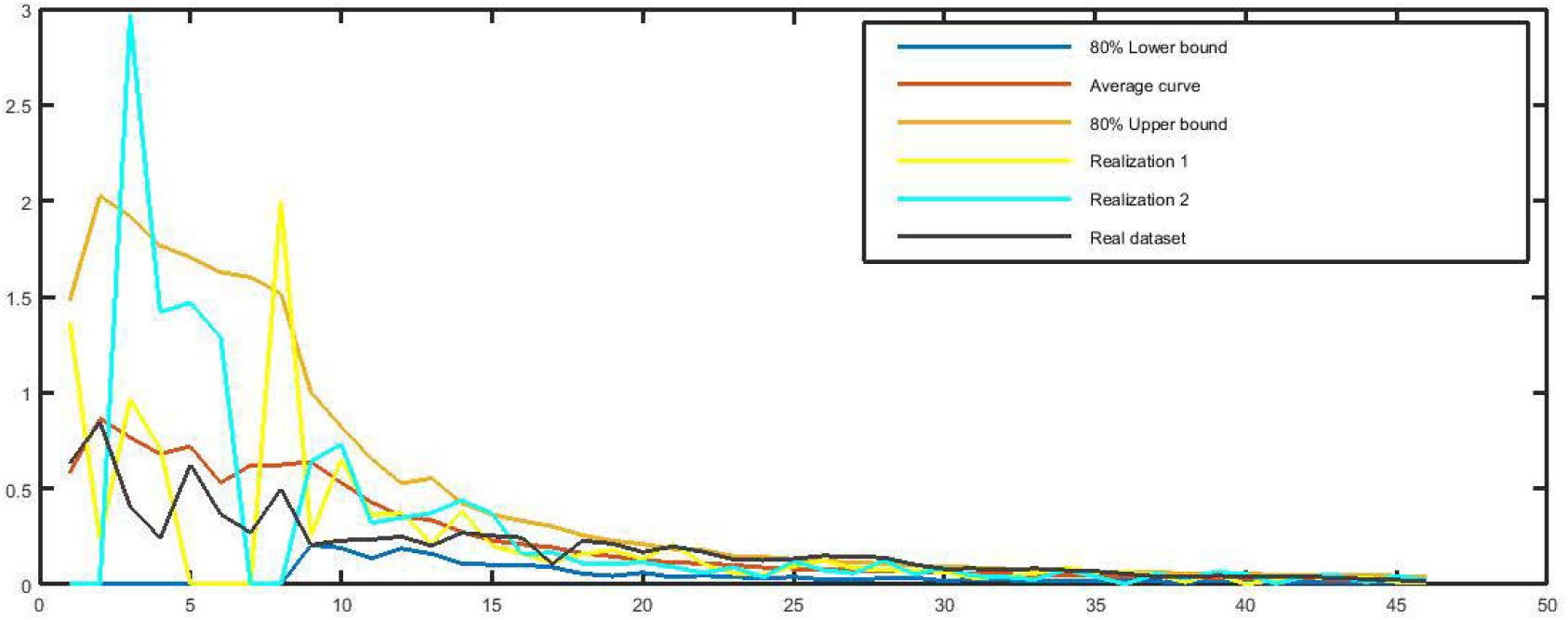
The relative increments of the number of confirmed cases, the 80% lower and upper bounds, the average curve, and two realizations of the model, 23.02.2020 – 08.04.2020.

Based on the obtained model, Plot 4 is obtained as a prediction of the variable of interest for 09.04.2020 to 18.05.2020. Based on this prediction, the relative increment decreases from 2.5% - 3% on 5-8 April to below 1% on 18 May. The model forecasts 80% confidence interval as [0.31% - 1.24%] with the mean equal to 0.78% for the relative increment on 18 May 2020 (Plot 4).

**Plot 4.**
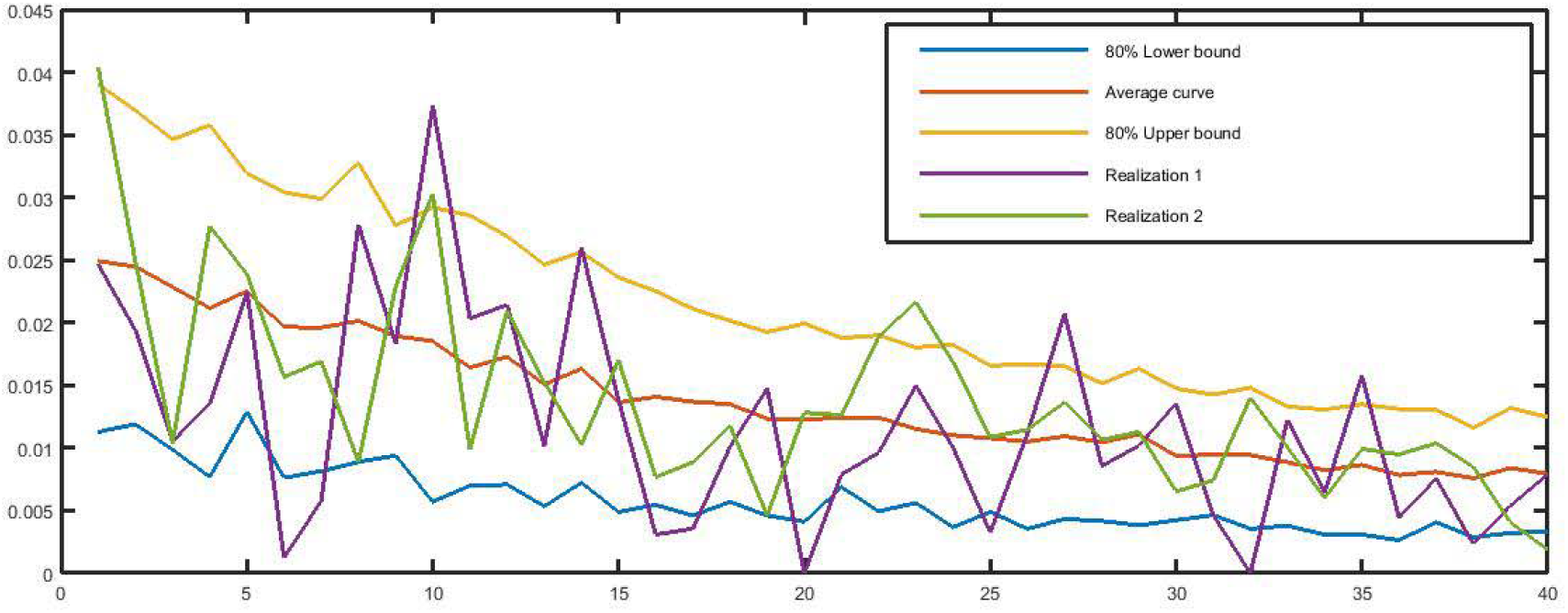
The forecast 80% lower and upper bounds, the average curve, and two realizations of the relative increments of the number of confirmed cases based on the model from 09.04.2020 to 18.05.2020.

Accordingly, we get Plot 5, as our prediction of the number of new confirmed cases daily from 09.04.2020 to 18.05.2020. Based on the model, the number of new confirmed cases falls from 3836 cases on 8 April to around 1900 cases on 18 May 2020. The 80% confidence interval of this variable is 520 – 4000.

**Plot 5.**
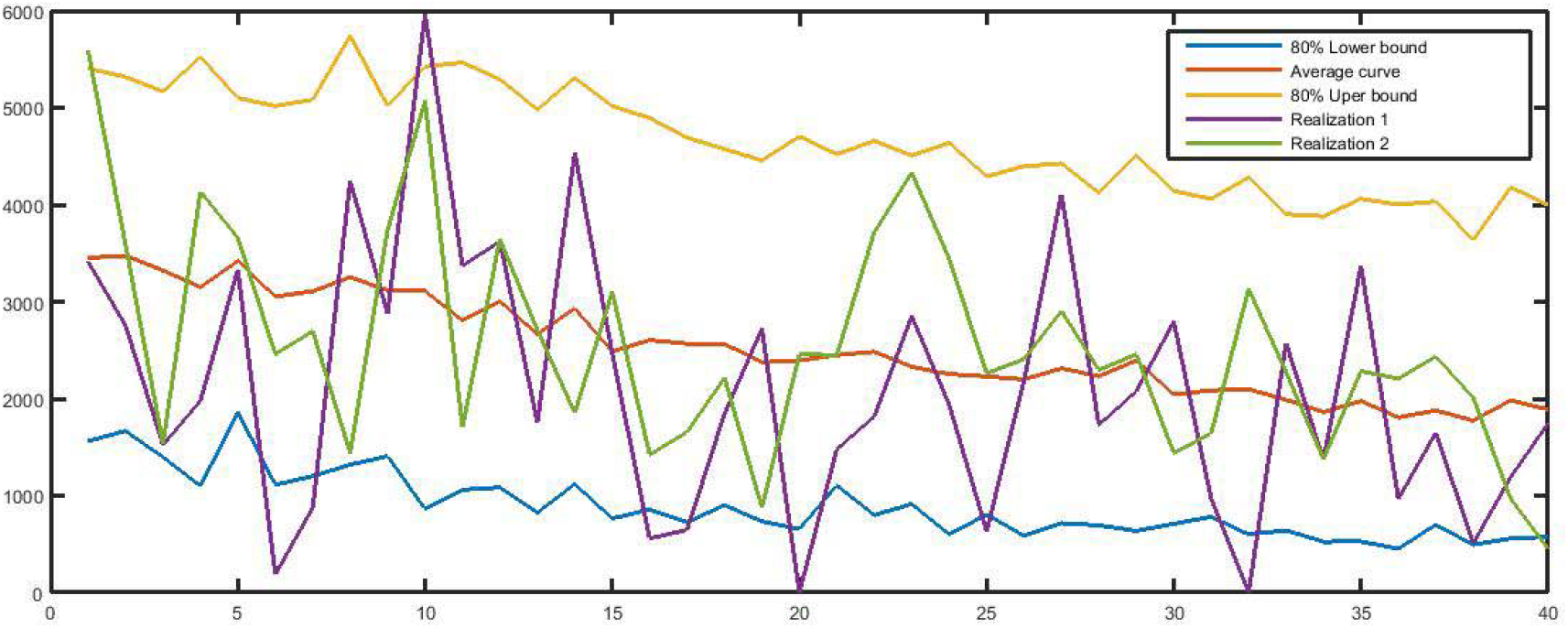
The forecast 80% lower and upper bounds, the average curve, and two realizations of the number of new daily confirmed cases based on the model from 09.04.2020 to 18.05.2020.

Finally, we obtain Plot 6 as the predicted cumulative confirmed cases for the studied period. We predicted that over the period 09 April to 18 May 2020, Italy will face 100000 new confirmed cases on average where considering around 140000 cases on 08 April, the cumulative number of confirmed cases gets 237635. It is noticeable that the 80% confidence interval for the predicted cumulative number of confirmed cases on 18 May 2020 is 226340 to 248417.

**Plot 6.**
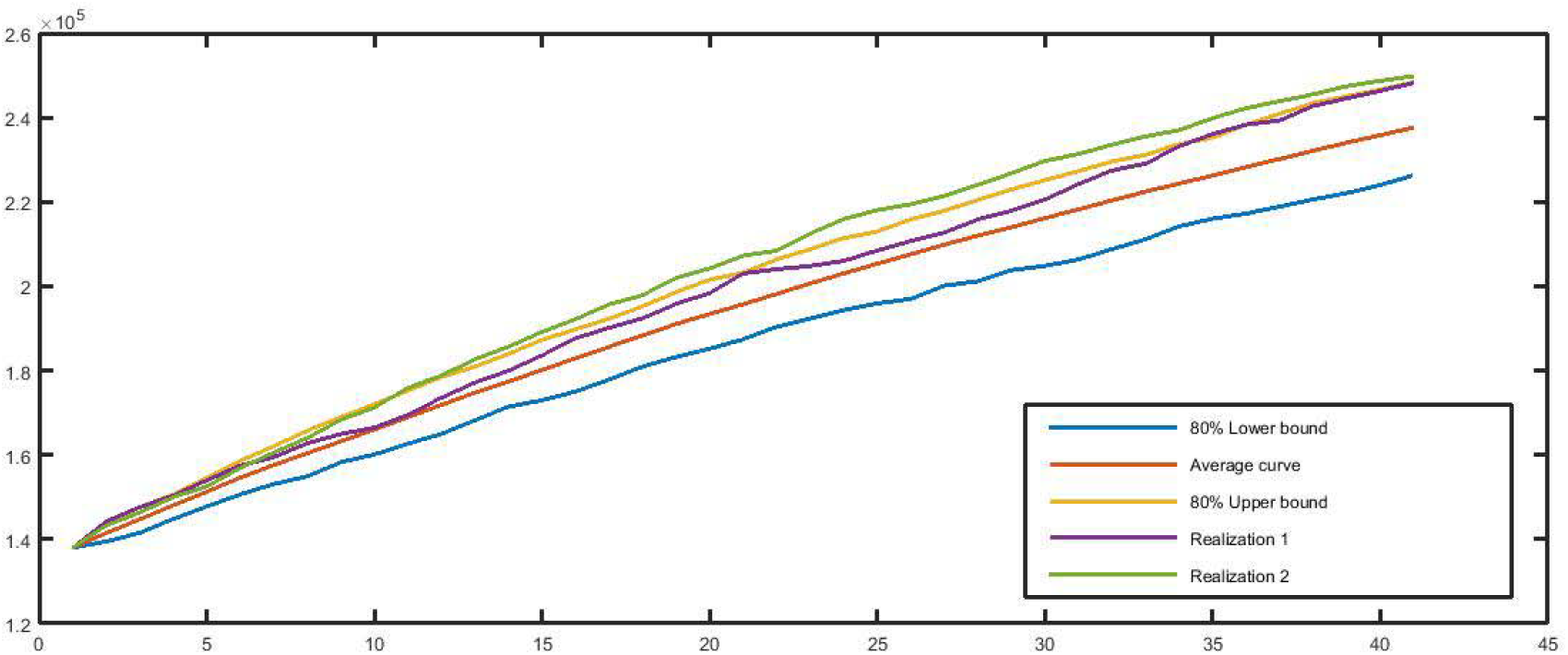
The forecast 80% lower and upper bounds, the average curve, and two realizations of the cumulative number of confirmed cases based on the model from 09.04.2020 to 18.05.2020.

## Discussions

Due to several characteristics such as highly social citizens, fair weather, densely populated cities, physically affectionate social interactions, and large elderly population, Italy has witnessed a severe COVID-19’s explosion. As of 8 April 2020, Italy, with 0.8% of the population of the world, has composed over 9% of all the confirmed cases and over 20% of the death toll from COVID-19.

Italy can be considered the first country to face the problem of the unpredictable and unprecedented explosion of the pandemic. Fortunately, these days, statistics establish that the country has managed to manage the disease to a large extent. Although it is a long path to fight to be over. However, it is very helpful to anticipate better for management and control of the disease. Accordingly, the present paper has predicted the number of confirmed cases in the next 40 days (April 9^th^ to May 18^th^). This prediction can be applicable in plans of policymakers regarding the management and hospitalization of the future infected people and the readiness of the staff involved, and inviting people to consider the health points.

According to our model, it is expected that by May 18^th^, 2020, the relative increment falls to the interval of 0.31% to 1.24% while on 8 April, it belonged to the interval 2.5% to 3%. Accordingly, It is expected that the new daily confirmed cases face a decreasing to around 1900 on average. Finally, our prediction established that the cumulative number of confirmed cases reaches around 240 K (with 80% confidence interval equal to [225 250] K by May 18^th^, 2020.

Finally, it is noticeable that the prediction is based on the assumption that the trend is stable, and there is no doubt that more strict adherence by people can lower the forecast, as well as non-compliance can lead to higher upward graphs.

## Data Availability

https://www.worldometers.info/coronavirus/coronavirus-cases/

## Contribution of the authors

BJ: Idea, Simulation, First manuscript

ATK: Final manuscript

SJZ: Literature, Simulation

MR: Revision

